# Research landscape of lymphovascular invasion in Oral Squamous Cell Carcinoma: A bibliometric analysis from 1994 to present

**DOI:** 10.1101/2023.02.27.23286490

**Authors:** Ankita Tandon, Kumari Sandhya, Narendra Nath Singh, Nikita Gulati

## Abstract

**Background:** The primary factor affecting tumor biology is neo-lymphangiogenesis in solid epithelial malignancies like OSCC. Determining the impact of lymphovascular invasion is critical in order to determine OSCC’s loco-regional, and global dissemination. Bibliometric landscapes are vital to learning about the most recent advancements in the aforementioned topic because the ongoing research in OSCC is multifaceted. This analysis can reveal the progressions that might modernize OSCC diagnosis and treatment.

**Objectives:** To study the relevance and effects of lymphovascular invasion in oral squamous cell carcinoma utilizing co-occurrence of keywords analysis and co-authorship analysis for the PubMed database.

**Methodology:** Cross-sectional bibliometric analysis of full-text PubMed articles from 1994 to the present using VOSviewer (Version 1.6.19) was performed. The keywords for the search of data included “Lymphovascular invasion in oral squamous cell carcinoma” using the Boolean operator (AND). The data obtained was analyzed for co-occurrence and co-authorship analysis using the VOSviewer standard protocol.

**Results:** The query revealed 296 searches in the PubMed database. Seven clusters were found with default colors in the representation of the entire term co-occurrence network, which also displayed a total link strength of 22262. The items were categorized into clusters based on their commonalities. The labels’ weights, as determined by Links and Occurrences, did not depend on one another, and the co-occurrence of keywords does not imply a causal association. In the item density visualization, item labels represented individual things. The number of items from a cluster that was close to the point was represented by the weight given to its color, which was formed by combining the colors of other clusters. A network of 57 authors who matched the search parameters was discovered by the co-authorship analysis. The network visualization map displayed three clusters with a total link strength of 184. The quantity of co-authorship relationships and the number of publications did not appear to be significantly correlated.

**Conclusion:** This investigation uncovered a sizable body of bibliometric data that emphasizes key trends and advancements in the aforementioned theme. The observed variances may be a result of the various objectives of the researchers and journals, who collaborate to provide the best possible literature dissemination.

**Graphic Abstract:** 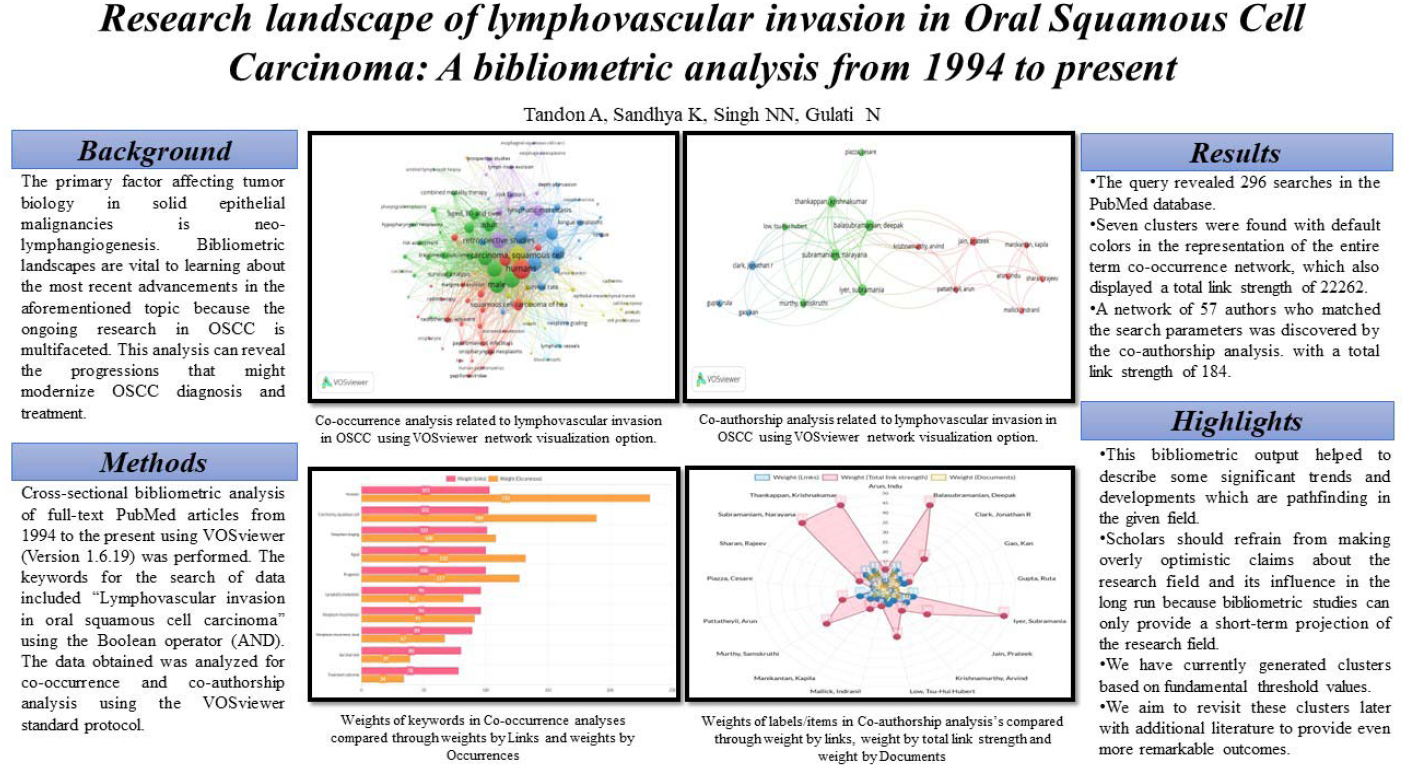

## Introduction

Oral Squamous Cell Carcinoma (OSCC) is a perennial major public health concern accounting for 90-95% of patients with this subtype of head and neck malignant diseases, followed by basal cell carcinomas, mesenchymal malignancies, hematologic tumors, and melanomas.^1^ The incidence of OSCC continues to rise and is anticipated to increase by 30% (that is, 1.08 million new cases annually) by 2030 (Global Cancer Observatory (GLOBOCAN).^2^

According to the classifications made by the World Health Organization’s International Agency for Research on Cancer (IARC), epidemiological studies have identified a wide range of risk factors for OSCC (WHO). The widespread occurrence of OSCC in Southeast Asia is associated with the usage of specific carcinogen-containing products, including alcohol consumption, smoking, environmental pollution exposure, and viral infections including HPV and EBV.^3^ In India, West Bengal has the highest incidence of oral cancer, whereas Kerala reports the lowest, according to epidemiological data.^4,5^

During the ongoing, intricate process known as lymphatic metastasis, cancer cells may migrate from the primary tumor site through the lymphatic system. Recent studies have demonstrated the importance of the tumor microenvironment (TME) in the growth, invasion, and metastasis of tumors.^6^ TME in OSCC is a complex and heterogeneous collection of tumor cells and stromal cells, including endothelial cells, cancer-associated fibroblasts (CAFs), and immune cells. Growth factors like VEGF, which are produced by CAFs and tumor cells, draw endothelial cells and encourage neovascularization, which increases the amount of oxygen and nutrients that reach the tumor.^3^ Compared to blood vessels, lymphatic capillaries are larger and do not have a continuous basal membrane, which makes it easier for cancer cells to invade lymphatic capillaries.^7^ OSCC causes neo-lymphangiogenesis, and most of these new lymphatic channels are located either intratumorally or peritumorally.^8^ The most recent National Comprehensive Cancer Network Guidelines for the treatment of head and neck squamous cell carcinomas included depth of invasion and extra nodal extension in the TNM staging of oral cancer. Although it can be difficult at times, lymphatic vessels are often difficult to identify from blood vessels.^9^ However, several studies in recent years have shown that it is possible to discriminate between blood vessels and lymphatic vessels using molecular markers, which has shifted research in this area in an effort to find more efficient treatment options for the cancer epidemic.

Although there is a wealth of information available regarding the multifactorial behaviour of OSCC, it has always been challenging to pinpoint the area of interest for study on the metastatic behaviour of cancer cells. As a result, bibliometric analysis, which examines research patterns using bibliographic data, can be used to track prior studies, predict future trends, and identify the current trend towards researching lymphovascular invasion in OSCC.

Therefore, the following bibliometric analysis was conducted to study the relevance and effects of lymphovascular invasion in OSCC utilising co-occurrence of keywords analysis and co-authorship analysis for the available PubMed database using the VOSviewer bibliometric software.

## Methodology

### Database and search strategy

Screening for “Lymphovascular invasion in oral squamous cell carcinoma” publications during a period from 1994 to present was done. Using the Boolean operator (AND), we performed a PubMed search (MEDLINE) for advanced search with keywords (“Lymphovascular”[All Fields] AND (“invasibility”[All Fields] OR “invasible”[All Fields] OR “invasion”[All Fields] OR “invasions”[All Fields] OR “invasive”[All Fields] OR “invasively”[All Fields] OR “invasiveness”[All Fields] OR “invasives”[All Fields] OR “invasivity”[All Fields]) AND (“squamous cell carcinoma of head and neck”[MeSH Terms] OR (“squamous”[All Fields] AND “cell”[All Fields] AND “carcinoma”[All Fields] AND “head”[All Fields] AND “neck”[All Fields]) OR “squamous cell carcinoma of head and neck”[All Fields] OR (“oral”[All Fields] AND “squamous”[All Fields] AND “cell”[All Fields] AND “carcinoma”[All Fields]) OR “oral squamous cell carcinoma”[All Fields])) AND (fft[Filter]) and text availability filter as “full text”.

### Methods for Co-occurrence analysis

The VOSviewer tool (Version 1.6.19) examined the co-occurrence of keywords “lymphovascular invasion in OSCC” patients using the data that was collected from the MEDLINE database. In order to illustrate the most prevalent keywords, a bibliographic map was created using the program’s full counting approach with “all keywords” as the unit of analysis and the minimum number of occurrences of a keyword set to five. The map’s elements were grouped into non-overlapping clusters and connected by lines with values. The stronger the relationship or co-occurrence between things, the higher the value.

On the VOSviewer, three different types of visualization were employed: the network visualization, which assigns keywords into clusters; the overlay visualization, which determines the average year of co-occurrence for each keyword; and the density visualization, which examines both item and cluster densities. Each point in the item density visualization had a color that denoted the density of items at that location, and things were represented by their labels in the item density visualization. Only when items had been assigned to clusters; the cluster density visualization was possible. The density of each cluster of items was displayed independently in the cluster density visualization as opposed to the item density visualization. The colors of numerous clusters were combined to form a point’s color in the cluster density display.

### Method for Co-authorship analysis

Between 1994 and present, at least three published documents with a total of no more than 25 authors were examined. The total co-authorship link with other writers was computed for each author. The network, overlay, and density visualizations which examined both item and cluster density were employed in the VOSviewer. The sum of the publications and co-authorship connections together represented the total number of publications per author.

## Results

### Co-occurrence analysis

All keyword co-occurrence network visualization showed seven clusters with a total of 104 items, 2965 links, and a total link strength of 22262. The first one, which was colored red and had 26 items, concentrated on the causes and symptoms of OSCC. The second cluster, shown by the blue tint, contained 24 items, mostly referring to OSCC treatment options and survival rates. 22 entries in the third cluster, which was colored green, highlighted the OSCC risk assessment characteristics and the lymphovascular invasion. The fourth cluster, which was comprised of 18 components and was highlighted in yellow, was centered on statistical models, metastasis, and OSCC tumor burden. The fifth cluster, which was highlighted in purple and contained ten items, focused on OSCC prognosis and lymph node features in metastasis. The sixth cluster, which was light blue in hue and contained two components, concentrated on the OSCC’s lymphovascular features. Only two elements made up the seventh cluster, which was orange and described the clinical traits of OSCC. **(Fig 1)**. The weights of labels (items) with predominance in searches across co-occurrence analyses are contrasted in **Graph 1** as weights (Links) and weights (Occurences). The graph emphasizes how labels, as measured by links and Occurrences, are independent of one another and that the keyword co-occurrence does not suggest a causal link association.

**Fig 1:**
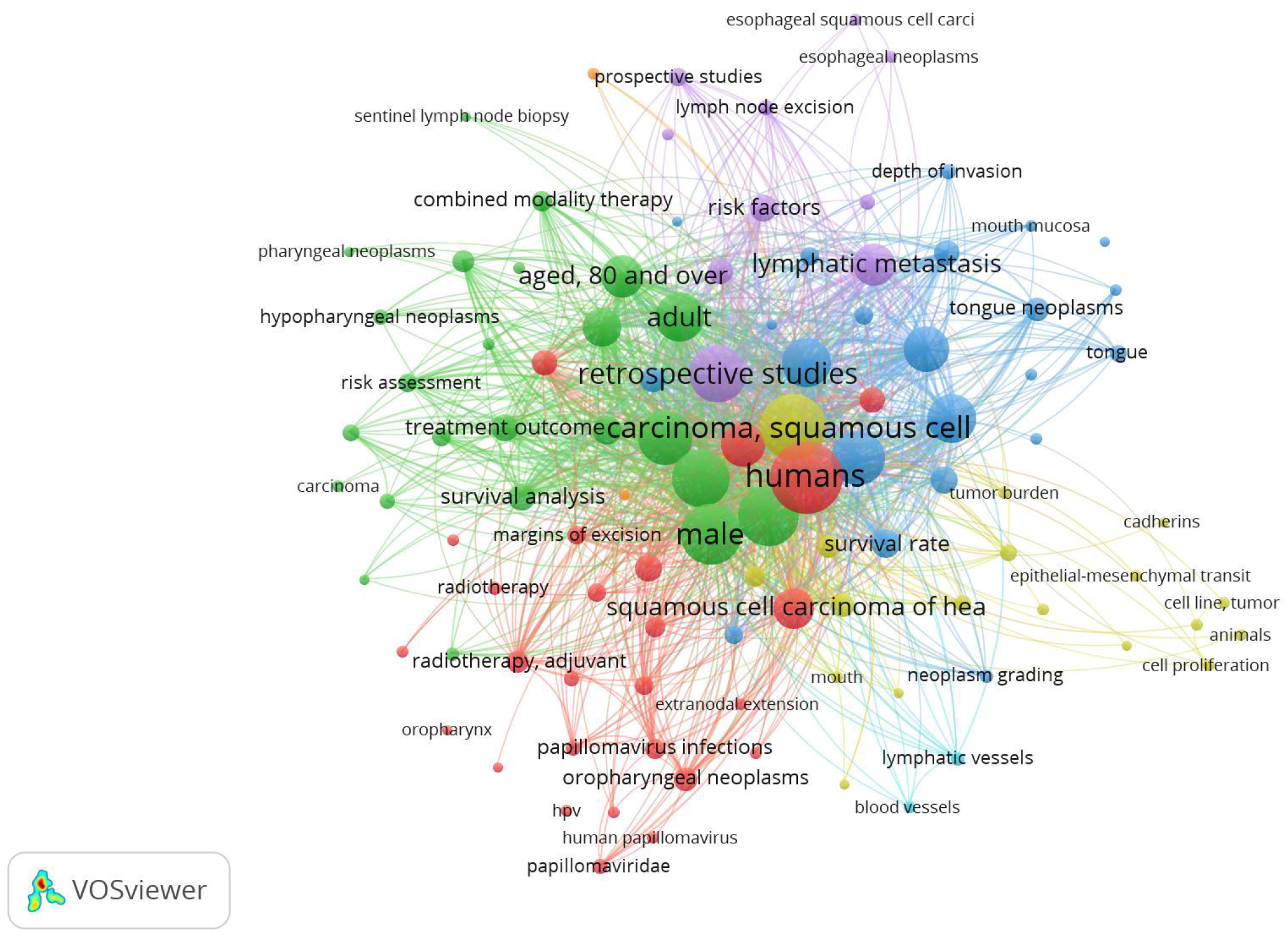
Co-occurrence analysis related to lymphovascular invasion in OSCC using VOS Viewer network visualization option.

Using the overlay visualization option on VOSviewer, the co-occurrence of all keywords showed that publications’ tendencies reflect a paradigm shift in their body of work. In the early research, the epidemiologic data and the clinical features of OSCC were emphasized the most. With the creation of more recent grading standards and risk assessment models, a minor increase in molecular evidence of risk factors was seen in the middle period between 1994 and the present. However, in the realm of OSCC research, the present literature is really focused on the aspects of the tumor microenvironment and cellular properties supported by histopathological and molecular data. **(Fig 2)**.

**Fig 2:**
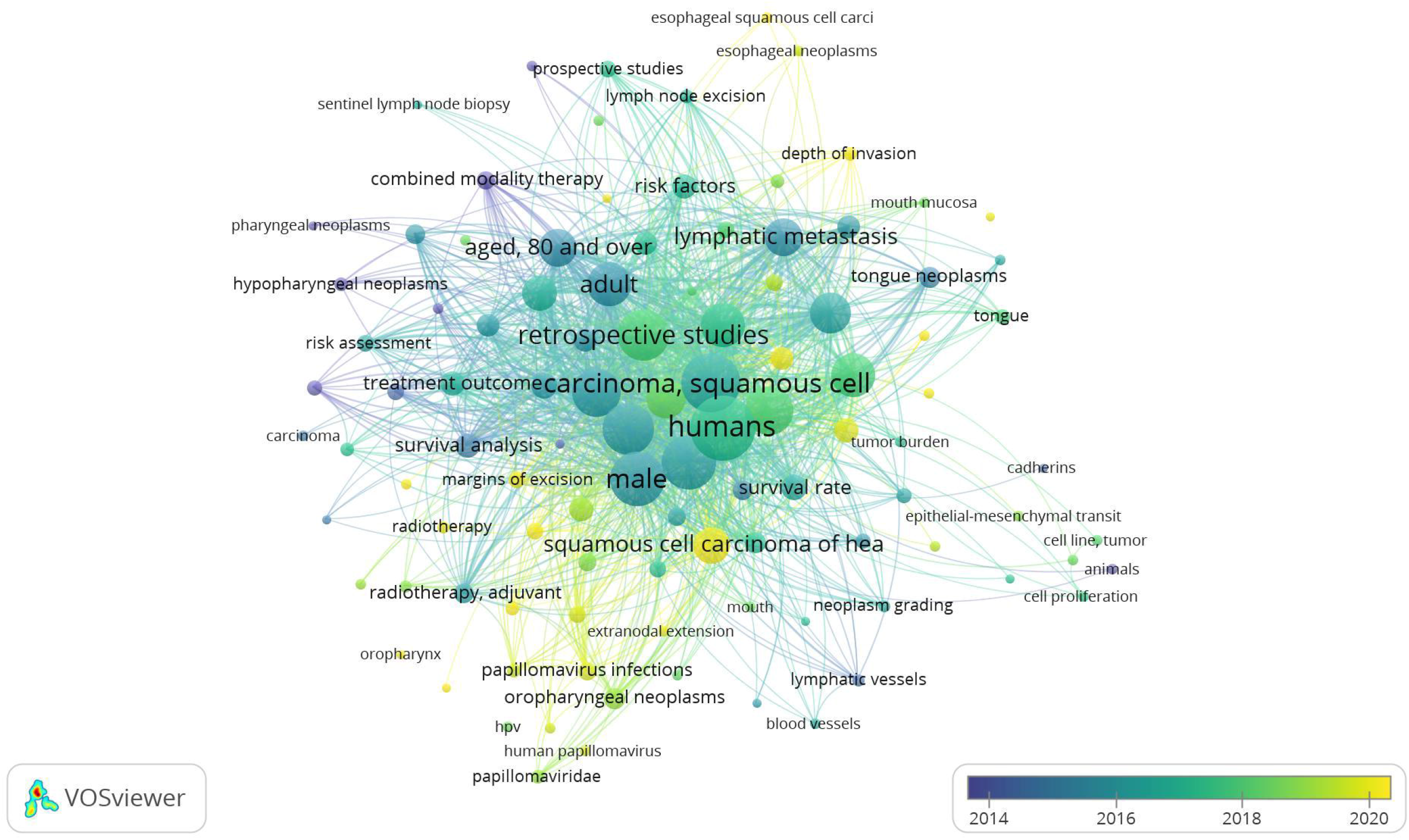
Co-occurrence analysis related to lymphovascular invasion in OSCC using VOS Viewer overlay visualization option (from Blue to Yellow color).

Using the density display feature of the VOSviewer, the co-occurrence of all keywords revealed item and cluster density. Item labels served as a representation of items in the item density visualization. Age, lymphatic metastasis, retrospective studies, men, etc. had more of these items nearby and hence had higher weights, making them look closer to yellow **(Fig 3a)**. Given that the items in the current analysis were assigned to clusters, the cluster density visualization was accessible. The weight assigned to a particular cluster’s color, which was created by combining the colors of other clusters, was based on how many items from that cluster were located close to the point **(Fig 3b)**.

**Fig 3:**
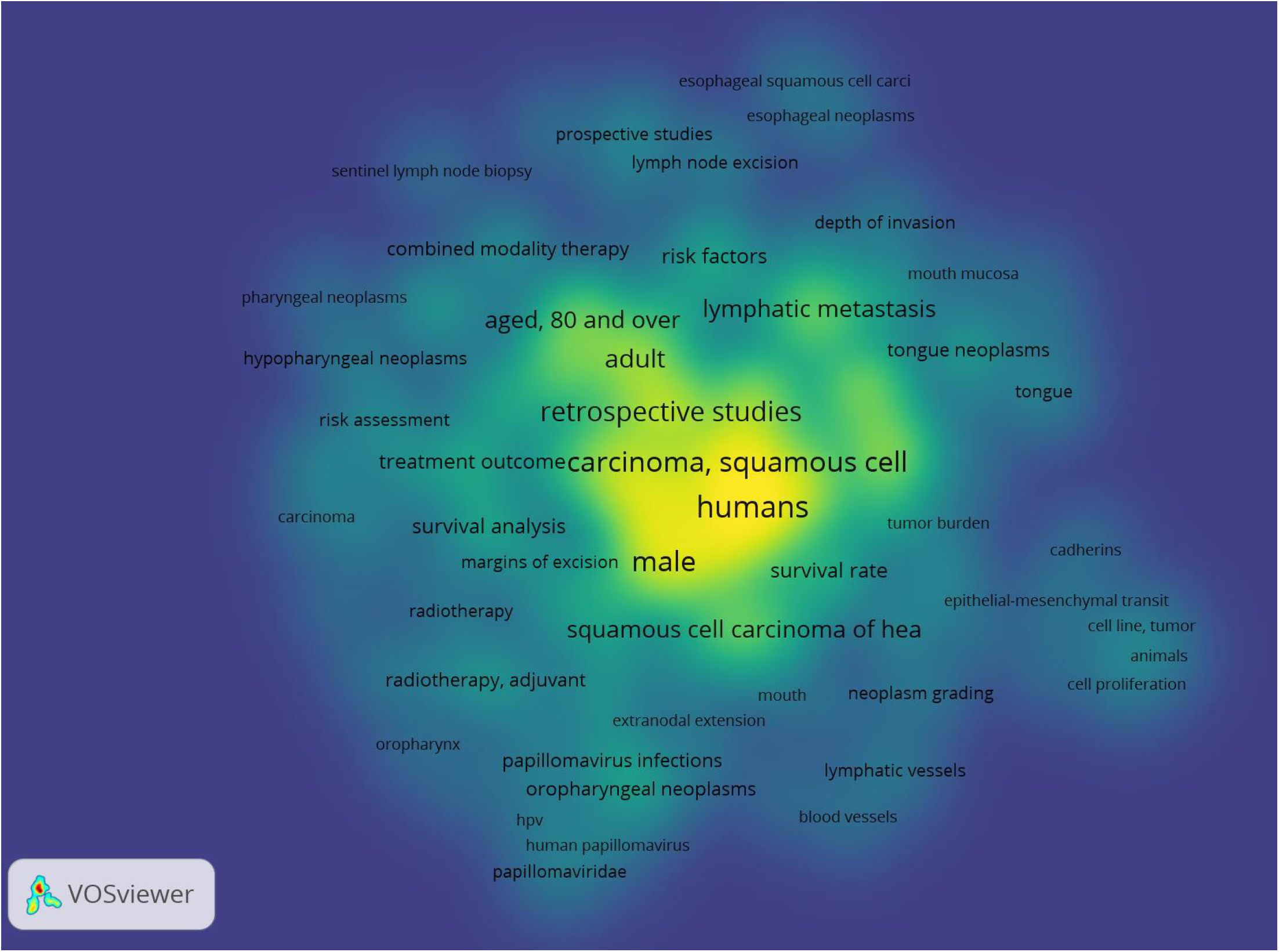
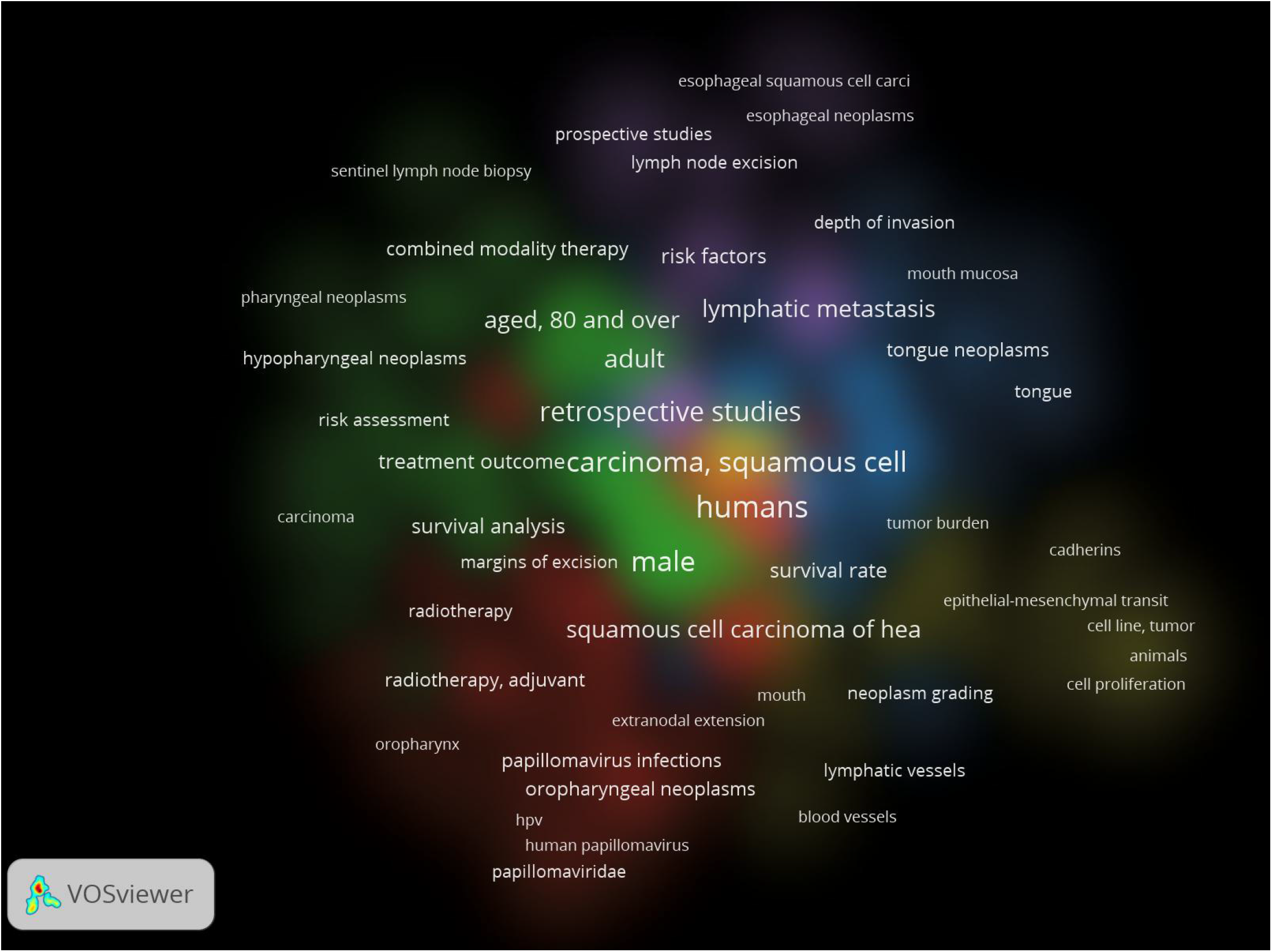
Co-occurrence analysis related to lymphovascular invasion in OSCC using VOS Viewer a) item density visualization and b) cluster density visualization options (mixture of colors).

### Co-authorship analysis

We looked at authors who had a minimum of three overall publications. 57 authors out of the total 1967 met the requirement. Three clusters were shown on the network visualization map, with 63 links and a total link strength of 184. **Fig. 4** shows the largest connected set, which was made up of 17 items. The total co-authorship link with other writers was computed for each author. It should be emphasized that the sum of publications and co-authorship relationships determines the overall number of publications per author. The 7 authors of cluster 2 (depicted in green) had a maximum number of documents to their credit. In **Graph 2**, the co-authorship analysis’s label (item) weight is displayed as weight (Links), weight (Total link strength) and weight (Documents). It was interesting to note that there was no clear correlation between the quantity of publications and the quantity of co-authorship linkages. The link strength between co-authors in one region of the world and those in another may be weaker even for those co-authors who have more publications (documents), and vice versa. According to the overlay scientific network representation **(Fig. 5)** based on all keyword searches, Indian researchers have recently dominated the OSCC research architype. These labels also served as representations of co-authorship in the item density visualization. According to document analysis, the following items from cluster 2 had higher weight labels: Thankappan, Krishnakumar; Balasubramanian, Deepak; Subramaniam, Narayana; and Iyer, Subramania **(Fig 6a)**. The same results were shown by the depiction of cluster density **(Fig. 6b)**.

**Fig 4:**
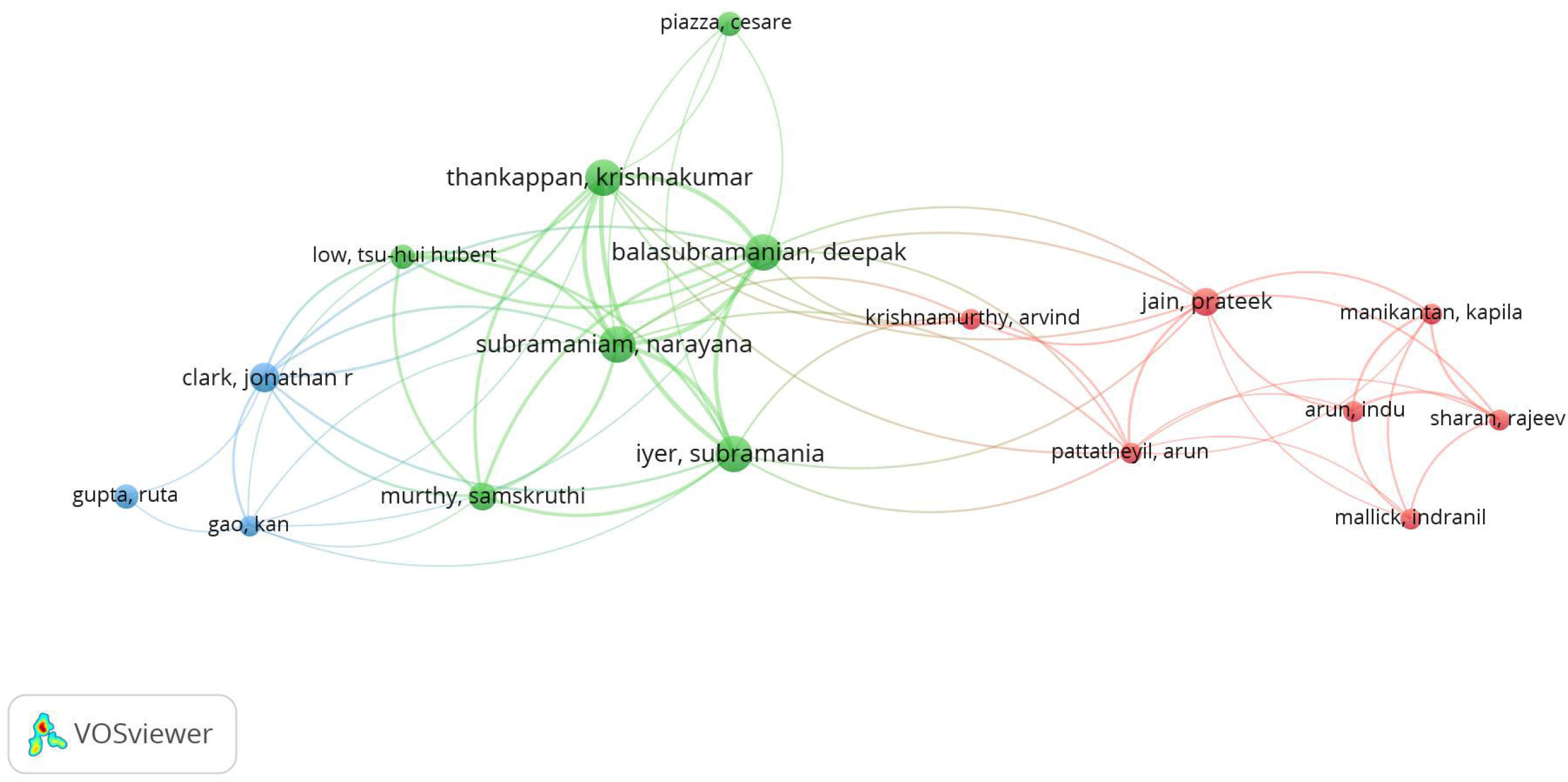
Co-authorship analysis related to lymphovascular invasion in OSCC using VOS Viewer network visualization option.

**Fig 5:**
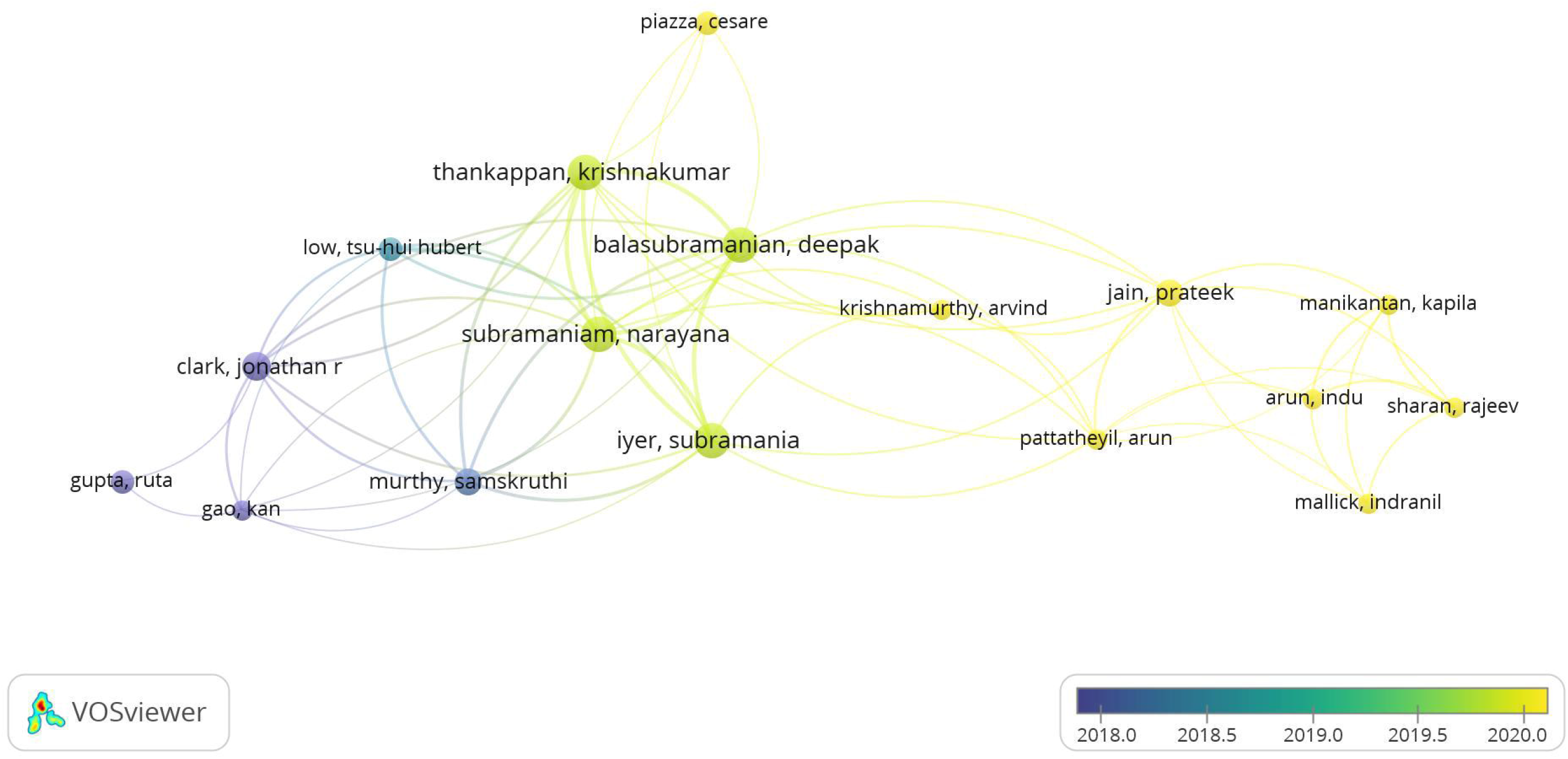
Co-authorship analysis related to lymphovascular invasion in OSCC using VOS Viewer overlay visualization option (from Blue to Yellow color).

**Fig 6:**
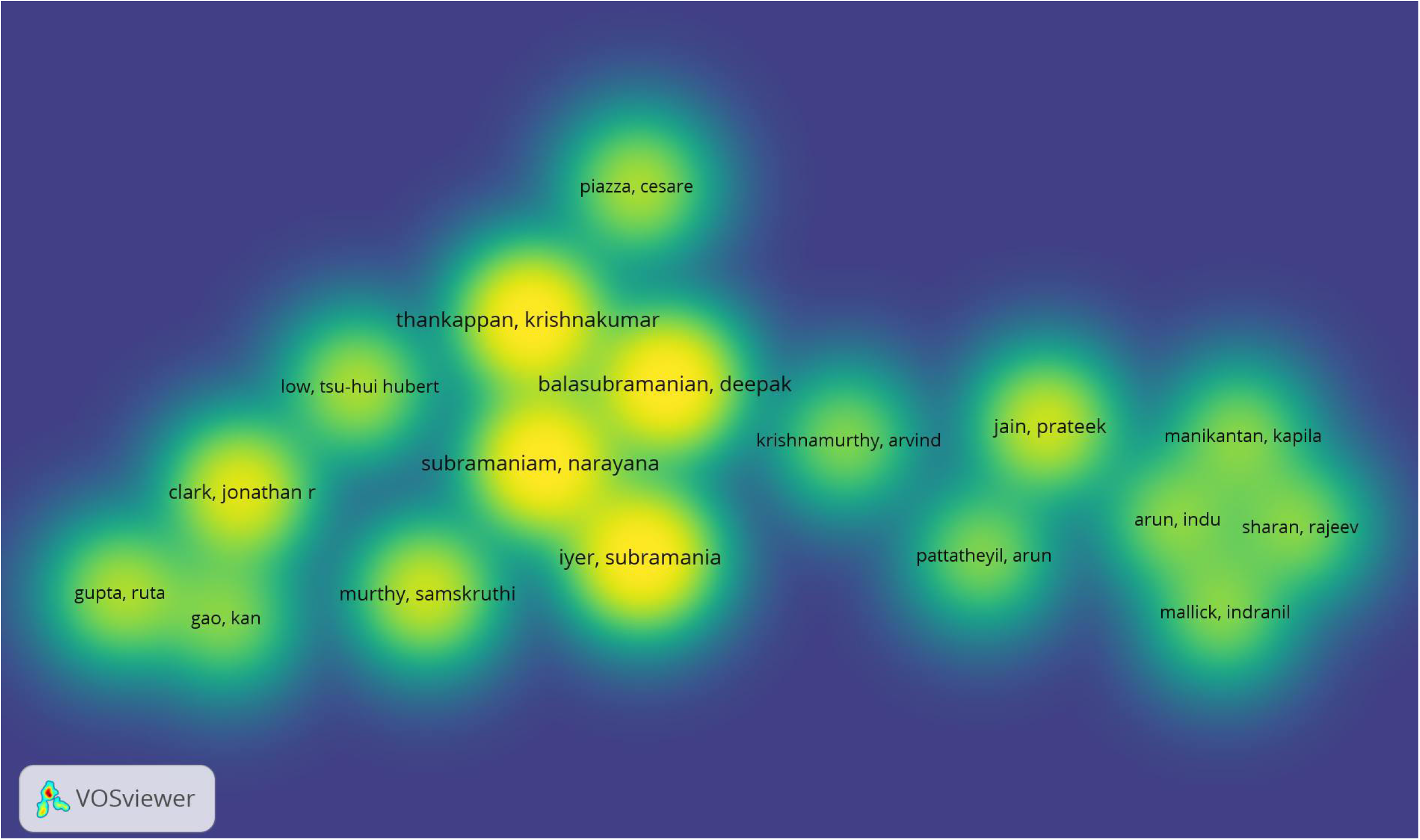
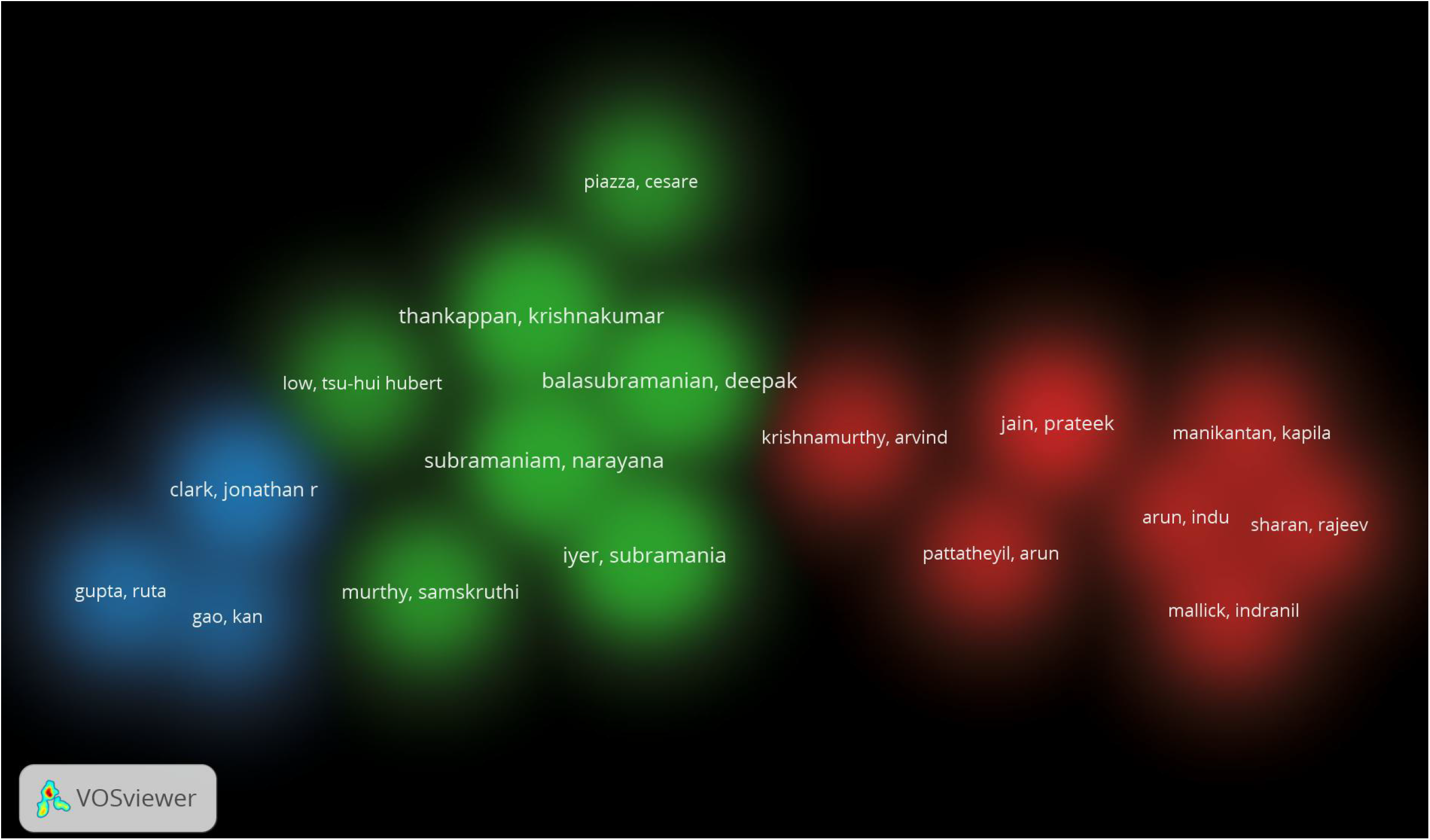
Co-authorship analysis related to lymphovascular invasion in OSCC using VOS Viewer a) item density visualization and b) cluster density visualization options (mixture of colors)

## Discussion

This bibliometric study presents a global research output of “lymphovascular invasion in Oral Squamous Cell Carcinoma”. We selected PubMed for data extraction as it is devoted for biomedical sciences, uses MeSH (Medical Subject Heading-a professional indexing tool) and is affiliated with several other National Library of Medicine (NLM) tools that can help to optimize analysis of biomedical subjects.^10^ The analysis was performed using VOSviewer due to its easy accessibility.

A core node with a higher number of direct connections to neighboring nodes exists in several of our networks. This representation of the major networks has the advantage that it makes it quick and easy to see who the key participants (items/labels) are and how they relate to one another. The dimensions, number, thickness, and number of connections that connect the nodes in a network helped to determine its relevance.^11^

When looking at the pool of available literature considering the keywords applied, the overall publication rate has increased substantially through decades, highlighting the rising importance of the topic and validating the need for thorough research in this arena. This positive trend reflects the increased focus of researchers in tumor microenvironment through high throughput molecular evidence. The seven clusters observed in co-occurrence analysis of our study, is descriptive of the fact that OSCC research is multi-dimensional and several etiopathologic factors govern the prognosis in OSCC **(Fig. 1-3)**. One such major component of tumor micro environment is lymphovascular invasion (LVI). It has been witnessed in the literature that amongst a variety of solid neoplasms, LVI is thought to be a crucial step in both local and distant metastasis.^12^ It is a pathologic process when tumor cells are found in specific endothelial-lined areas, such as lymphatic or blood vessels.^13^ LVI has recently been added as another prognostic factor for OSCC in the eighth AJCC staging system, which supports the growing body of evidence showing it is a poor prognostic factor for oral cancer patients^14^ with a higher chance of loco-regional recurrence, cervical lymph node metastasis, and overall worse prognosis. LVI can therefore be used as a parameter to determine the aggressiveness and to choose patients in the future for more focused and aggressive treatment.^15^ In our study, **Graph 1** shows the top ten labels (items) according to their weight from bibliometric search wherein the labels (items) displayed significant variation in their link strength, indicating the non-uniformity in OSCC research’s global focus.

Scientific collaboration is a crucial research topic and an essential part of today’s academic landscapes across disciplines and research sectors.^11^ According to the evaluation of co-authorship analysis, the strongest network of researchers consisted of 17 items with a notable degree of variation in their body of work. The most reliable criterion was the weight assessment of labels by document analysis (number of publications by each author) **(Graph 2)**. The majority of these authors discussed lymphovascular invasion in OSCC in their presentations on the tumor microenvironment **(Fig. 4-6). Table 1** lists the authors in order of their label weights as determined by the examination of their individual documents. Their works were published in numerous journals with impact factors ranging from 2.136 to 13.608. The weight of labels obtained from document or link assessment can also be evaluated, and it can be found that it is not always proportional. From this, it could be deduced that there is a poor collaboration among some authors and that authors with higher publications tend to collaborate with wide range of other authors and vice versa. **Fig. 7** provides a complete analysis of the distribution of co-authorship among institutes and nations through infographics.

**Table 1:**
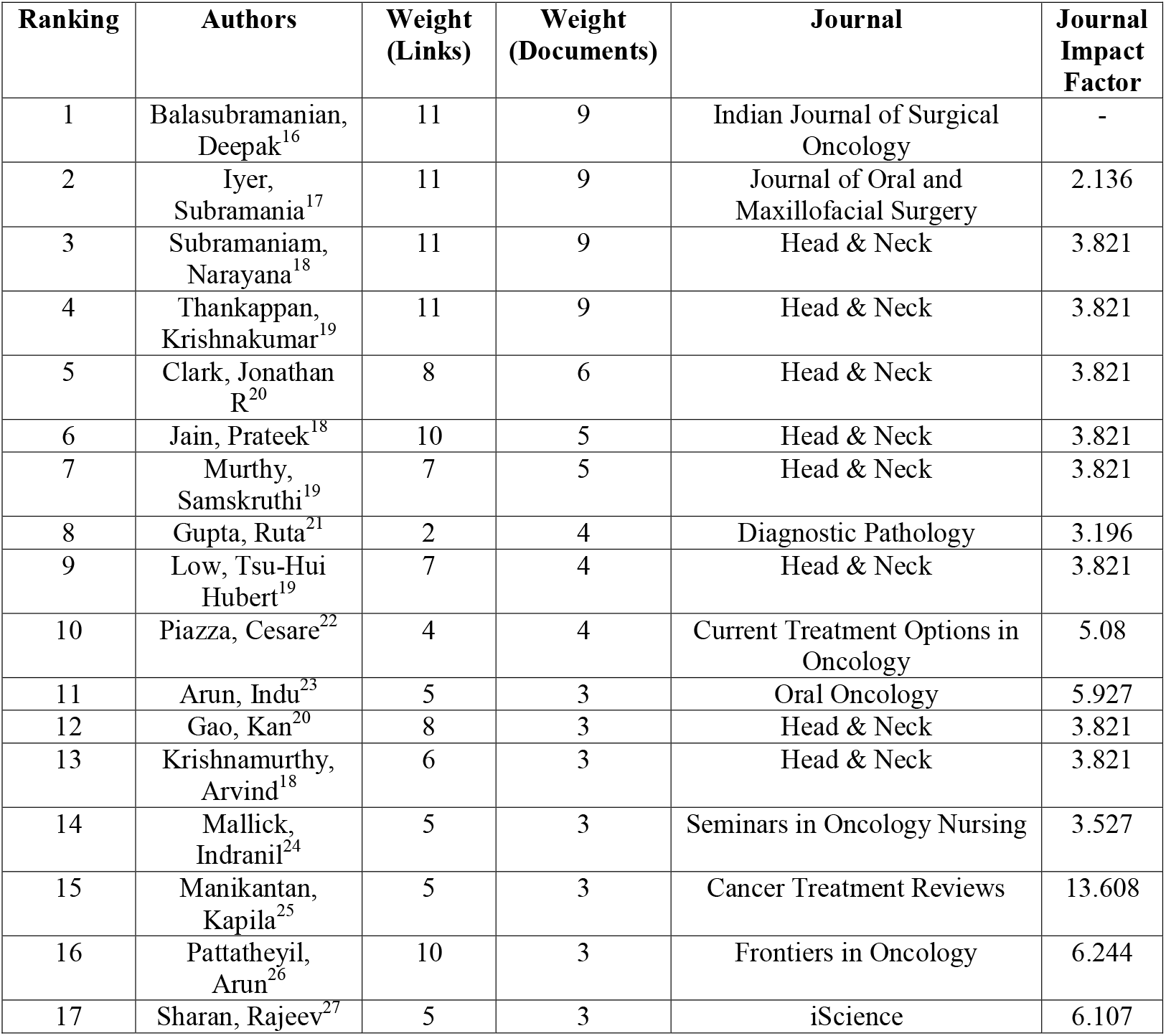
Author ranking and their associated journal analysis.

**Fig 7:**
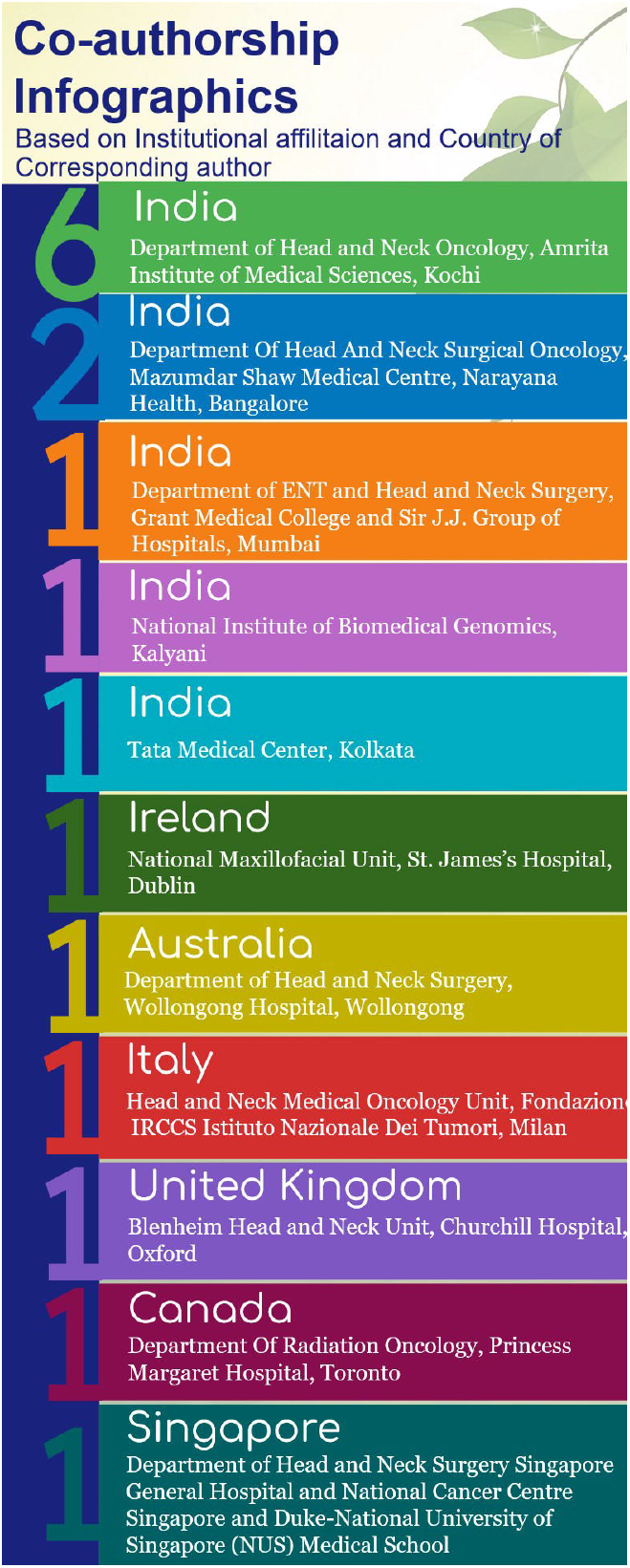
Co-authorship infographics representing association amongst institutes and nations (the numbers display per institutional contributions in a country)

## Limitations

This was the first effort to analyze the impact of research on lymphovascular invasion in OSCC. There are certain restrictions to our bibliometric analysis, though. For instance, we limited our analysis to the PubMed database. Due to their subscription requirements, the databases provided by Web of Science and Scopus could not be included. Also, data exported from PubMed could not be used for identifying citation, bibliographic coupling, and co-citation links between items. Hence, when working with PubMed data, some options in the Create Map wizard of VOSviewer were not available. Additionally, to maintain sample homogeneity, only English-language literature was included.

## Conclusion (Highlights of the present analysis)

- This bibliometric output helped to describe some significant trends and developments which are pathfinding in the given field and can promote interdisciplinary collaboration among scholars, enabling them to unite and delve deeper into the aforementioned theme.
- Grouping studies based on topics and article characteristics might help to pinpoint the main areas where the exploration is needed.
- Scholars should refrain from making overly optimistic claims about the research field and its influence in the long run because bibliometric studies can only provide a short-term projection of the research field.
- We have currently generated clusters based on fundamental threshold values.
- We aim to revisit these clusters later with additional literature to provide even more remarkable outcomes.

## Data Availability

All data produced in the present study are available upon reasonable request to the authors

## Figure legends

**Graph 1:**
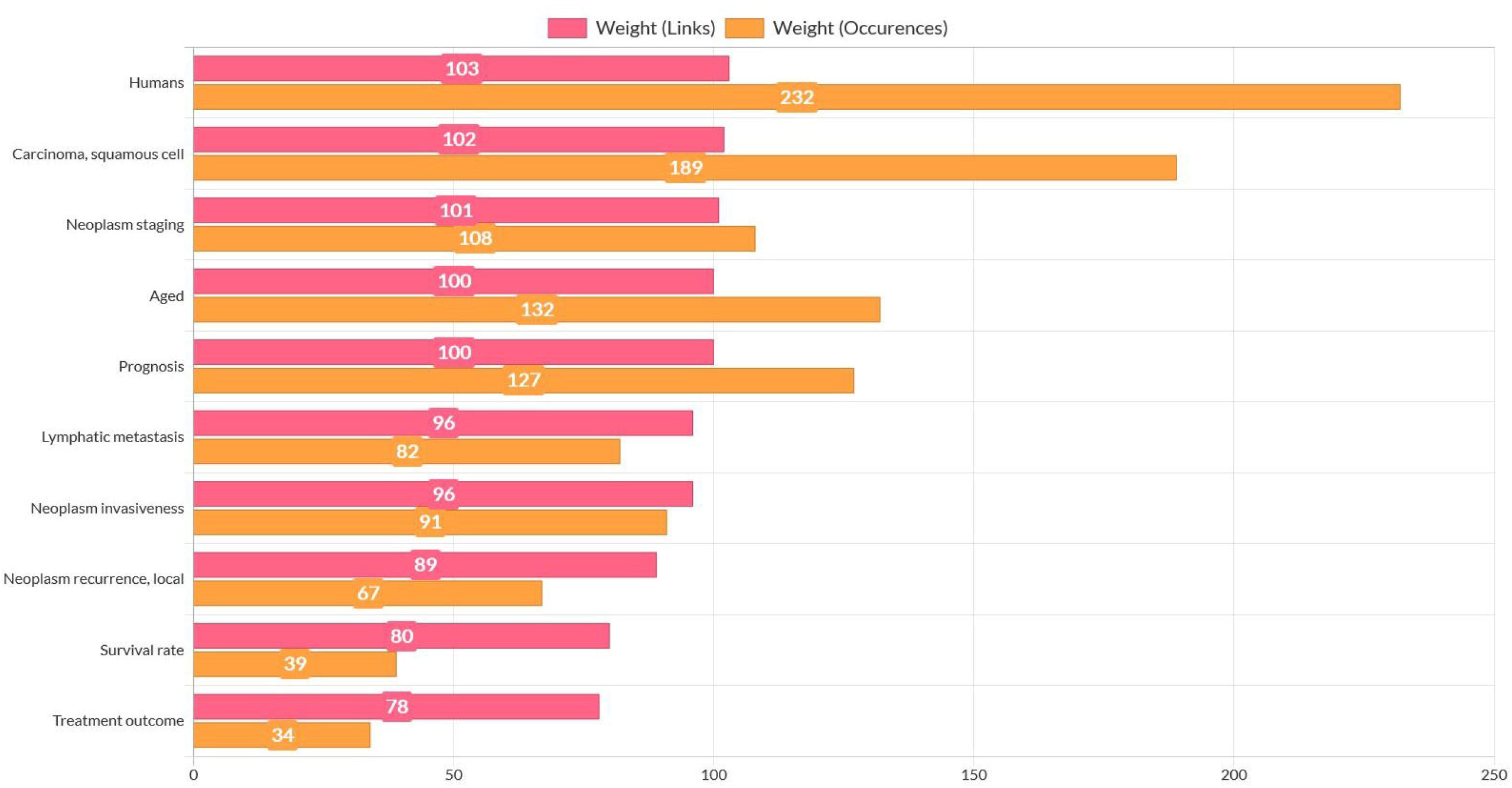
Weights of keywords in Co-occurrence analyses compared through weights by Links and weights by Occurrences

**Graph 2:**
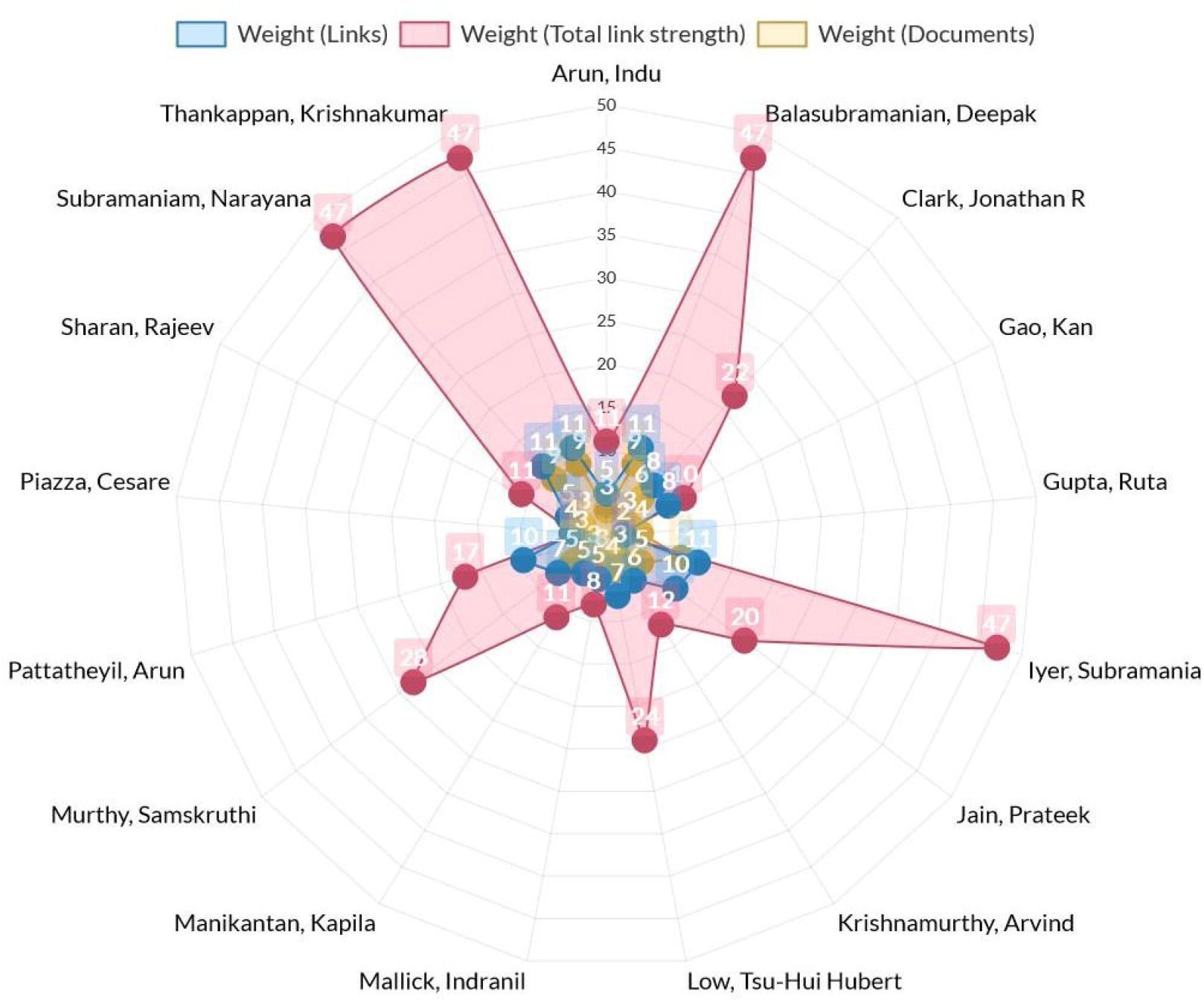
Weights of labels/items in Co-authorship analysis’s compared through weight by links, weight by total link strength and weight by Documents

